# Association between Haloperidol use and Risk of Rheumatoid Arthritis

**DOI:** 10.1101/2023.09.11.23295367

**Authors:** Vidya L. Ambati, Tammy H. Cummings, Praveen Yerramothu, Joseph Nguyen, S. Scott Sutton, Brian C. Werner, Joseph Magagnoli

## Abstract

Haloperidol is an anti-psychotic used for the treatment of schizophrenia or Tourette disorder. Here we report, by studying three large administrative health insurance databases, that haloperidol use is associated with a reduced risk of developing rheumatoid arthritis. A meta-analysis revealed a 31% reduced hazard of incident rheumatoid arthritis among individuals with schizophrenia or Tourette disorder treated with haloperidol compared to those treated with other anti-psychotic drugs. These findings suggest a potential benefit of haloperidol in rheumatoid arthritis and provide a rationale for randomized controlled trials to provide causal insights.

## MAIN

An uncontrolled case-series reported a benefit of haloperidol in rheumatoid arthritis (RA) (1). A study of the FDA Adverse Events Reporting System reported that among RA patients there were fewer users of anti-psychotic drugs, including haloperidol (2). This suggests RA is associated with a reduced risk of anti-psychotic use but not that anti-psychotics reduce the risk of RA. In a small employee insurance database in Japan, this study also reported an inverse temporal relationship between RA and anti-psychotic drugs, including haloperidol. Limitations of these studies include small size, unverified and incomplete records, lack of correction for sociodemographic or clinical confounders, changing time-trends in prescriptions, and selective loss to follow-up. Therefore, we sought to undertake a robust pharmacoepidemiologic study of whether haloperidol, an FDA-approved drug for the treatment of schizophrenia or Tourette disorder, affects the risk of incident RA in three large nationwide health insurance databases that comprise most of the United States population.

We studied three health insurance databases: PearlDiver Mariner (2010-2021), IBM Marketscan Research Databases (2006-2020), and Veterans Administration (VA) Health database (2001-2021). Inclusion criteria were at least 6 months follow-up, at least 18 years of age, schizophrenia or Tourette disorder diagnosed on 2 separate occasions, and treatment with anti-psychotics. RA prior to diagnosis of schizophrenia or Tourette disorder was exclusionary. Disease claims were identified by ICD-9-CM and ICD-10-CM codes. Drug exposure was determined by prescriptions for generic or brand versions. Time from first prescription of anti-psychotics to diagnosis of RA was the dependent variable. Sensitivity analyses were performed using diagnosis of schizophrenia or Tourette disorder as the index date and flagging treatment with anti-psychotics within 60 days of schizophrenia or Tourette disorder diagnosis. Patients were censored when they developed RA, left the plan, died, or switched between study medication groups. Cox proportional hazards regression analyses were performed to estimate the hazard of RA in relation to haloperidol use. We performed propensity score matching (3) to assemble cohorts with similar baseline characteristics, reducing possible bias in estimating treatment effects. Additionally, to control for any residual covariate imbalance, we adjusted for RA-associated confounders: age, sex, smoking, body mass index, Charlson comorbidity index, and database entry year. Fine-Gray subdistribution hazard ratios for competing risk of mortality, corrected for covariates, were calculated for the VA, which contains mortality data. We performed inverse-variance weighted meta-analyses to estimate the combined hazard ratio (HR) and 95% confidence intervals (CI) using a random-effects model, computed prediction intervals, and Kaplan-Meier survival analyses.

The adjusted Cox proportional hazards regression models in the propensity-score-matched populations showed a protective association of haloperidol use against incident RA in each of the three databases (Figure 1). The meta-analyses identified a protective effect of haloperidol in the main (pooled aHR = 0.69; 95% CI, 0.61, 0.79; P = 0.0001) and sensitivity analyses (pooled adjusted hazard ratio (aHR) = 0.69; 95% CI, 0.58, 0.81; P < 0.0001) (Figure 1). The 95% prediction intervals for the hazard ratio, which provide insights into the range of outcomes in a hypothetical future study (4), were < 1.0 in both analyses (Figure 1), implying a future clinical trial would be estimated to have a >95% probability of identifying a protective effect. Subdistribution hazard ratios in the VA revealed a protective effect (main analysis: aHR = 0.52; 95% CI, 0.27, 0.98; P = 0.0002; sensitivity analysis: aHR = 0.48; 95% CI, 0.30, 0.76; P = 0.0002). Kaplan-Meier survival analyses revealed that haloperidol use was associated with a lower risk of incident RA in all three databases in both analyses (Figure 2).

**Figure 1.**
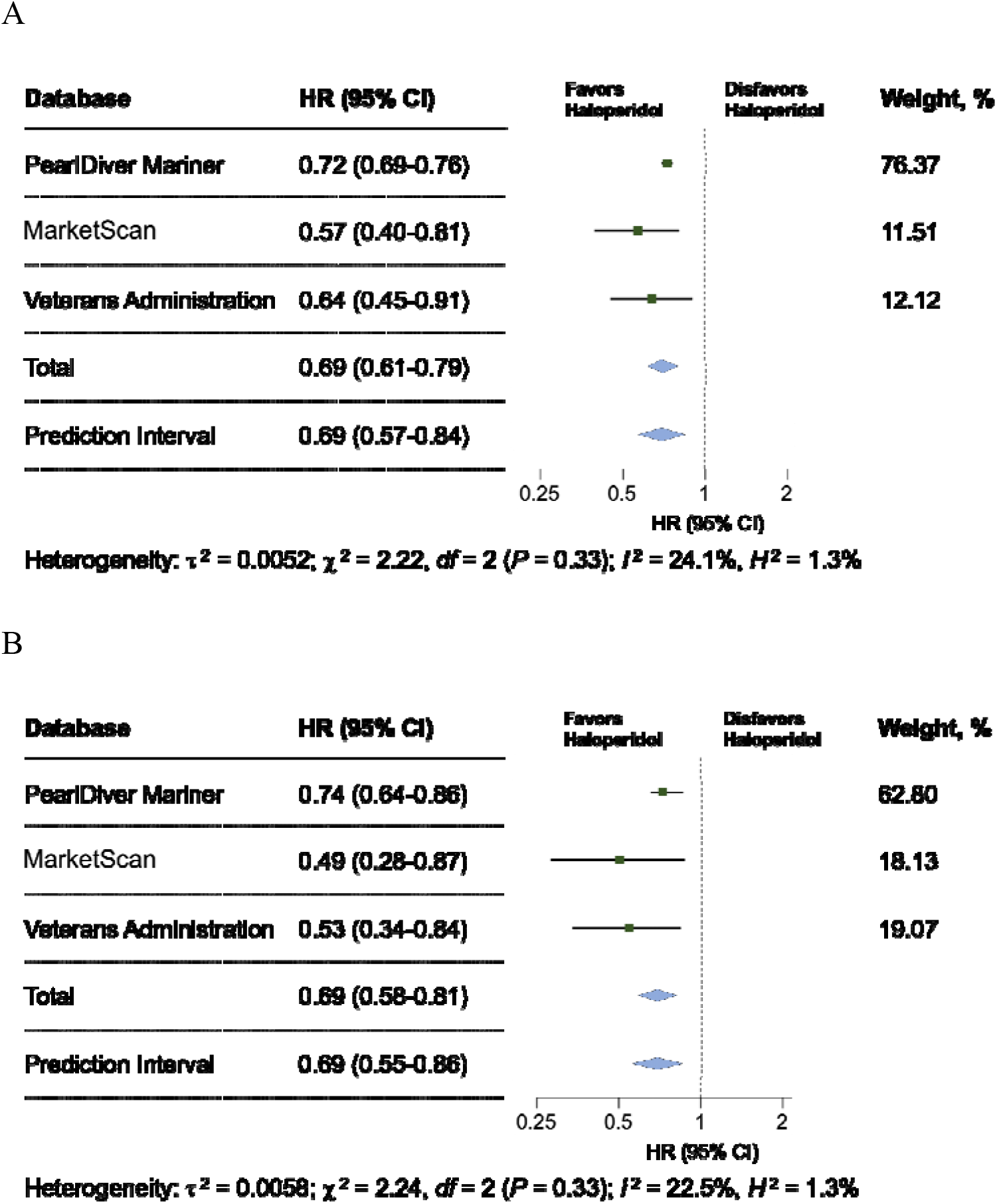
Forest Plots of Incident Rheumatoid Arthritis for Haloperidol Exposure. Hazard ratios (HR) for developing rheumatoid arthritis derived from propensity score-matched models adjusted for the confounding variables listed in Methods were estimated separately for database. (A) Main analysis: index date is first prescription of anti-psychotic. (B) Sensitivity analysis: index date is initial diagnosis of schizophrenia or Tourette disorder. Adjusted hazard ratios (green squares) along with 95% confidence intervals (CI) as whiskers. The blue diamond shows the pooled estimate of the adjusted hazard ratio and the 95% confidence intervals for meta-analyses using an inverse-variance-weighted random-effects model. A blue diamond also shows the prediction interval. P values (χ^2^ test) and weights assigned to each database per the random-effects model are reported.

**Figure 2.**
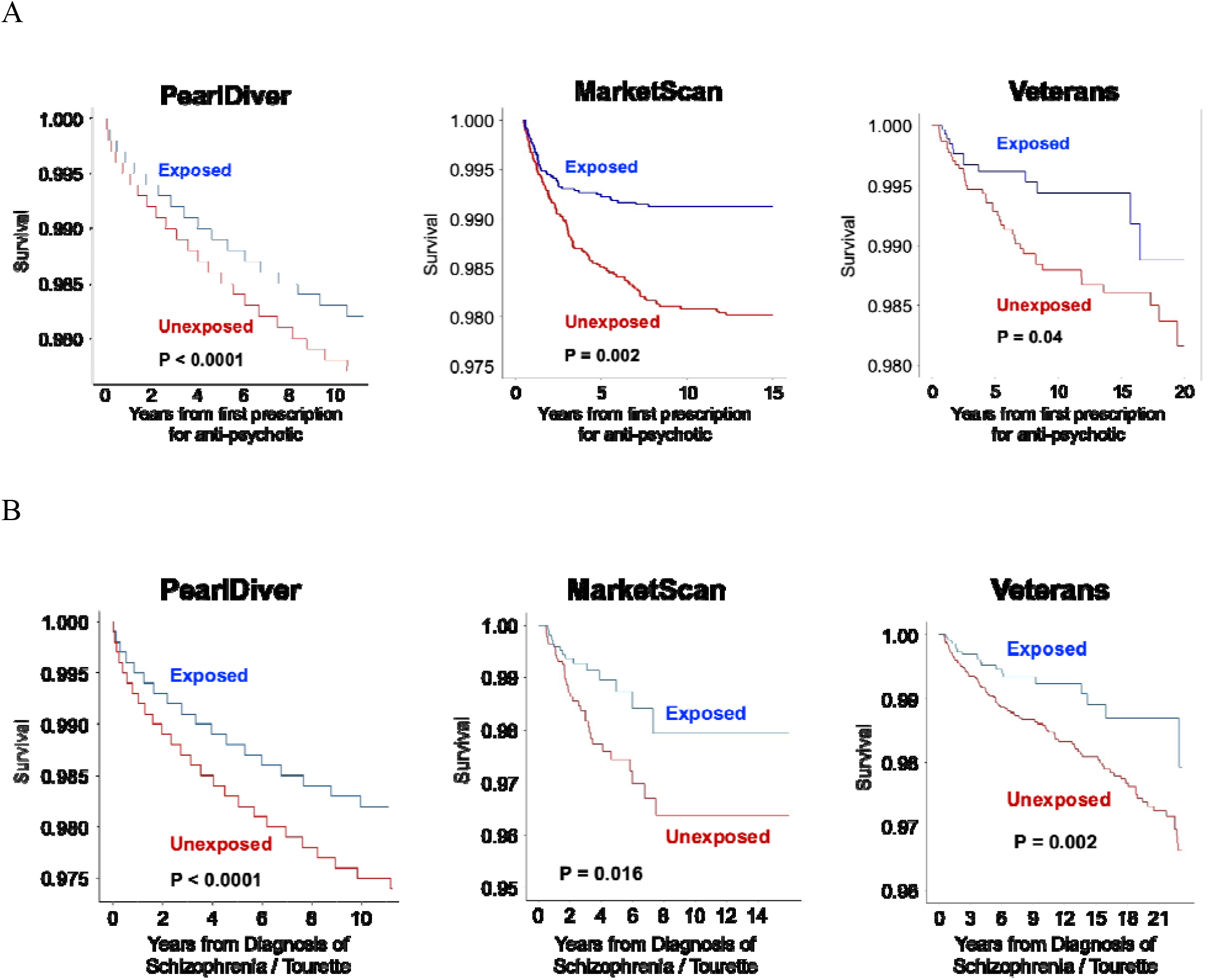
Kaplan-Meier Estimates of Incident Rheumatoid Arthritis for Haloperidol Exposure and Non-exposure. (A) Main analysis: index date is first prescription of anti-psychotic. (B) Sensitivity analysis: index date is initial diagnosis of schizophrenia or Tourette disorder. P values by log-rank test.

We identified a significant association between haloperidol use and incident RA. Strengths of our study include replication in three cohorts that comprise most United States adults, adjustment for confounders, and propensity score matching, which simulates randomization and increases its validity. As this retrospective study was not randomized, there could be residual confounding or selection bias. These findings suggest a potential benefit of haloperidol in RA and provide a rationale for randomized controlled trials to provide causal insights.

## AUTHOR CONTRIBUTIONS

**Vidya Ambati:** Conceptualization; data curation; formal analysis; funding acquisition; methodology; writing – original draft; writing – review and editing.

**Tammy H. Cummings:** Funding acquisition; writing – review and editing.

**Praveen Yerramothu:** Data curation; Writing – review and editing.

**Joseph Nguyen:** Data curation; writing – review and editing.

**S. Scott Sutton:** Data curation; funding acquisition; methodology; writing – original draft; writing – review and editing.

**Brian C. Werner:** Conceptualization; data curation; funding acquisition; methodology; supervision; writing – original draft; writing – review and editing.

**Joseph Magagnoli:** Conceptualization; data curation; formal analysis; funding acquisition; methodology; supervision; writing – original draft; writing – review and editing;.

All authors have read and approved the final manuscript.

## DATA AVAILABILITY STATEMENT

All data supporting the findings of this study are available within the paper. Analyses of the Veterans Health Administration Database were performed using data within the US Department of Veterans Affairs secure research environment, the VA Informatics and Computing Infrastructure (VINCI). The other health insurance datasets are subject to licensing agreements and privacy restrictions. Researchers interested in accessing aggregate and individual data are encouraged to make direct enquiries to the corresponding author and should note they may also need to approach PearlDiver and IBM MarketScan for access to data from these sources.

## ETHICS STATEMENT

PearlDiver Mariner and IBM Marketscan Research Databases contained de-identified data deemed exempt by the University of Virginia Institutional Review Board (IRB). Veterans Administration (VA) Health database data analysis was compliant with Department of Veterans Affairs requirements and received IRB and Research and Development Approval.

